# Digital exclusion predicts worse mental health among adolescents during COVID-19

**DOI:** 10.1101/2021.11.25.21266853

**Authors:** Thomas E. Metherell, Sakshi Ghai, Ethan M. McCormick, Tamsin J. Ford, Amy Orben

**Affiliations:** Department of Psychology, University of Cambridge, Cambridge, UK; MRC Cognition and Brain Sciences Unit, Cambridge, UK; Donders Institute, Radboud University Medical Center, Nijmegen, NL; Department of Psychiatry, University of Cambridge, Cambridge, UK

## Abstract

**Background:** Social isolation is strongly associated with poor mental health. The COVID-19 pandemic and ensuing social restrictions disrupted young people’s social interactions and resulted in several periods during which school closures necessitated online learning. We hypothesise that digitally excluded young people would demonstrate greater deterioration in their mental health than their digitally connected peers during this time.

**Methods:** We analysed representative mental health data from a sample of UK 10–15-year-olds (*N* = 1387); Understanding Society collected the Strengths and Difficulties Questionnaire in 2017-19 and thrice during the pandemic (July 2020, November 2020 and March 2021). We employed cross-sectional methods and longitudinal latent growth curve modelling to describe trajectories of adolescent mental health for participants with and without access to a computer or a good internet connection for schoolwork.

**Outcomes:** Adolescent mental health had a quadratic trajectory during the COVID-19 pandemic, with the highest mean Total Difficulties score around December 2020. The worsening and recovery of mental health during the pandemic was greatly pronounced among those without access to a computer, although we did not find evidence for a similar effect among those without a good internet connection.

**Interpretation:** Digital exclusion, as indicated by lack of access to a computer, is a tractable risk factor that likely compounds other adversities facing children and young people during periods of social isolation.

**Funding:** British Psychological Society; School of the Biological Sciences, University of Cambridge; NIHR Applied Research Centre; Medical Research Council; Economic and Social Research Council; and Emmanuel College, University of Cambridge.

---

Since the onset of the COVID-19 pandemic, populations around the world have experienced a noted decrease in mental health, with evidence for rising levels of anxiety, depression, and psychological distresses.^1^ Large-scale disruptions to work, education, leisure, and social activities, as well as additional pandemic stresses and healthcare problems, make such a development unsurprising. Nevertheless, specific concerns have been raised about the mental health impacts of the pandemic on adolescent populations. Adolescence represents a vulnerable period for the development of mental health disorders,^2^ and these challenges can have long-lasting consequences into adulthood.^3,4^ The mental health of children and adolescents in the United Kingdom was already deteriorating before the pandemic, highlighted by increases in anxiety, depression and self-harm.^5–7^ Since the onset of the pandemic, however, the incidence of probable mental health conditions in this age group has risen further from 10·8% in 2017 to 16% in July 2020,^8^ a trend mirrored in other studies that found deteriorating mental health in adolescents both in the UK^9^ and internationally.^10^

One of the most prominent disruptions to adolescent life during the COVID-19 pandemic has been the closure of schools and the increase in online schooling.^11,12^ While school closures caused educational disruptions experienced by most adolescents,^13^ their impact was not felt equally. For those adolescents who were digitally excluded, (i.e. lacking the computer or internet access needed to successfully partake in online-only education) educational disruptions were much greater.^14^ For example, in a UK sample, 30% of school students from middle-class homes reported taking part in live or recorded school lessons daily, while only 16% of students from working-class homes reported doing so.^14^ Prior research has shown that educational disruption can negatively impact adolescent mental health,^15^ and this may have resulted in negative impacts on adolescents’ mental health outcomes during the pandemic.

In addition to school closures, COVID-19 and the subsequent lockdown measures brought with them a curtailing of general social contact and widespread social disruption. At times when in-person peer interaction was cut to a minimum, online and digital forms of interaction with peers (e.g., through video games or social media) might have helped buffer some of these social disruptions.^16,17^ Lack of access to the technologies necessary to support online interactions could have therefore led to negative mental health consequences, especially as adolescent cognitive, biological and social development makes them more sensitive to limitations in social contact and decreases in peer interaction.^18^ Digital exclusion therefore had the potential to further exacerbate the mental health impacts experienced by adolescents due to technology’s role in allowing adolescents to not just partake in educational activities, but also to stay involved in social life.

In this study we use *Understanding Society*, a large longitudinal panel survey from the United Kingdom to test whether the mental health trajectories for adolescents who were digitally excluded during the pandemic differed from those of their digitally included counterparts. While this question has not been systematically investigated, research has found that adolescent mental health was not uniformly impacted by the pandemic,^19^ and that more socioeconomically disadvantaged children and young people showed worse mental health.^20^ We therefore first examine trends in mental health across the pandemic using latent growth curve modelling, and then test whether these models differ for those digitally excluded compared with those with digital access.

## Methods

### Study design and participants

In this study, we analysed data from the UK Household Longitudinal Study (Understanding Society),^21,22^ a longitudinal survey of around 40 000 UK households, with data collected annually from adults and adolescents since January 2009. At age 10, household members are first included in the survey via a paper self-completion ‘youth questionnaire’; participants migrate to the adult questionnaire at age 16. In this study we used youth questionnaire data from wave 9 as the baseline for our analysis (with invitations being issued between January 2017 and December 2018) – this being the last main study wave for which youth mental health data are available. During COVID-19, additional youth surveys were administered bimonthly between April and July 2020, and every four months between September 2020 and March 2021. Here we use data from COVID-19 waves 4, 6, and 8, dictated by when the mental health questionnaire was issued as part of the survey. 2862 unique youth questionnaires were returned in main study wave 9 (2017-19), 1411 in COVID-19 wave 4 (July 2020), 1432 in COVID-19 wave 6 (November 2020), and 1388 in COVID-19 wave 8 (March 2021). We employed longitudinal weights as calculated and described by the data custodians for COVID-19 wave 8.^23^

The University of Essex Ethics Committee approved all data collection for the Understanding Society main study and innovation panel waves, including asking consent for all data linkages except to health records.

### Procedures

To measure mental health, we used the Strengths & Difficulties Questionnaire (SDQ)^24^ (paper, self-completed and returned in a sealed envelope), which was included in the youth questionnaire in odd-numbered main study waves (until wave 9 in 2017-19) and even-numbered COVID-19 waves (from wave 4 in July 2020). The SDQ comprises 25 items that assess common childhood psychological difficulties through a series of positively and negatively phrased statements, which are rated as ‘not true’, ‘somewhat true’ and ‘certainly true’. Items are subsequently scored from 0 to 2 as appropriate, so that a high score indicates greater difficulty. The SDQ has five subscales: *hyperactivity/inattention, prosocial behaviour, emotional, conduct and peer relationship problems*. The total of all but the prosocial scale are summed to provide a total difficulties score that ranges from 0 to 40, which we used to measure adolescent mental health^24^ in 2017-19 (main study) and July 2020, November 2020 and March 2021 (COVID-19 survey).

To group participants by digital inclusion, we used the following question, included in the COVID-19 wave 6 (November 2020) youth questionnaire (paper, self-complete): *“Which of these things do you have at home to help you do your school work?”* The question included the response options “*Access to a… computer” and “…good internet connection”*.

The sociodemographic variables used as control variables in this study were sex (male vs female), age (on 15^th^ August 2020 in whole years), ethnicity (dichotomised, white vs non-white; these three derived from youth questionnaire responses) and household income (monthly, averaged across the four waves of interest; derived from the individual responses of adults in the same household).

### Statistical analysis

Participants with a calculated youth longitudinal weight^23^ for COVID-19 wave 8 (March 2021), and at least one recorded SDQ Total Difficulties score across the four waves, were included in this analysis (*N* = 1387). When grouping by computer or internet access, participants with missing data for the digital inclusion question (39·7%) were excluded (*n* = 836). Analyses were conducted in R using the R package *lavaan*^25^ and graphs produced using the R package *ggplot2* among others. Sampling probability weights were accounted for in analyses with a sufficiently large sample using the R packages *survey*^26^ and *lavaan*.*survey*.^27^

We analysed the data by fitting latent growth curve models (LGMs)^28^ to the Total Difficulties scores, including multi-group models to model disparate mental health trajectories of adolescents with and without computer and internet access. Full information maximum likelihood estimation^29,30^ was employed to minimise the biasing impact of missing data. We took a sequential model-selection approach with two main components: 1) establishing the proper functional form of the mental health trajectory experienced by adolescents during the COVID-19 pandemic, and 2) establishing the appropriate level of measurement invariance^31^ between digitally included and excluded groups. Model comparisons were performed via pairwise scaled *χ* ^2^ tests of goodness of fit^25^ (i.e. likelihood ratio test; LRT). To test for functional form, we first compared an intercept-only model to a linear model, and then a quadratic model. For the multi-group models, additional pairwise comparisons were conducted to test model fit with equality constraints between groups, including factor variances, factor covariances, and the Total Difficulties residual variances. In so doing we established whether there is significant evidence for differences in model parameters between the two groups in each case. Mis-specified models (e.g., models with negative variances or a non-positive definite covariance matrix) were discarded during the model building process. To probe the robustness of our analyses, we also conducted a sensitivity check where those participants without longitudinal weights were not excluded, and refitted models with sociodemographic covariates grouping by both computer and good internet connection access (see appendix tables A12-15 and figures A1-2).

## Results

Before fitting our longitudinal models, we cleaned our data and excluded participants without a longitudinal weight or any mental health scores. 1388 participants had an assigned longitudinal weight^23^ for COVID-19 wave 8, of which 1 (0·07%) had no Total Difficulties scores and so was excluded, leaving 1387 (656 [47·3%] male, 731 [52·7%] female) adolescents to be included in our analyses. Of these, 638 (46·0%) had a Total Difficulties score in main study wave 9, 818 (59·0%) in COVID-19 wave 4, 836 (60·3%) in COVID-19 wave 6 and 1386 (99·9%) in COVID-19 wave 8. Full characteristics of participants with missing responses are provided in the appendix (table A1). Of the 1387 included participants, 836 (60·3%) had a valid response to the digital inclusion question. Table 1 shows the sociodemographic characteristics of participants in relation to their responses to the digital inclusion question.

**Table 1.** Digital inclusion characteristics according to key sociodemographic variables (ethnicity data suppressed due to low numbers to protect the identities of participants)

| Group | Response type |  |  | Has a computer/has access to a good internet connection |  |
| --- | --- | --- | --- | --- | --- |
|  | All | Available | Unavailable | Yes/Yes | At least one no |
| **Total** | 1387 | 836 | 551 | 714 | 122 |
| **Sex** |  |  |  |  |  |
| Male | 656 | 386 | 270 | 343 | 43 |
| Female | 731 | 450 | 281 | 371 | 79 |
| Unavailable | 0 | 0 | 0 | 0 | 0 |
| **Birth year** |  |  |  |  |  |
| 2004–2007 | 781 | 481 | 300 | 419 | 62 |
| 2008–2011 | 606 | 355 | 251 | 295 | 60 |
| Unavailable | 0 | 0 | 0 | 0 | 0 |
| **Ethnicity** |  |  |  |  |  |
| White | 1005 | .. | .. | .. | .. |
| Non-White | 367 | .. | .. | .. | .. |
| Unavailable | 15 | .. | .. | .. | .. |
| **Mean household income (x, annual)** |  |  |  |  |  |
| $x < £40,000$ | 378 | 250 | 128 | 205 | 45 |
| $£40,000 \leq x$ | 407 | 260 | 147 | 228 | 32 |
| Unavailable | 602 | 326 | 276 | 281 | 45 |

In the sensitivity check (see methods), we also included those participants without a longitudinal weight (see appendix tables A12-15 and figures A1-2), incorporating a total of 1422 valid responses to the digital inclusion question.

### Overall profile of mental health

To understand the general trend in mental health over time, we first examined the raw SDQ data. The mean Total Difficulties score was 10·7 (out of a maximum 40) in main study wave 9 (2017-19) and then peaked at 11·4 (out of a maximum 40) in COVID-19 wave 6 before declining to 11·1 in COVID-19 wave 8 (Figure 1). This shows that there are small changes in mental health across the pandemic.

**Figure 1.**
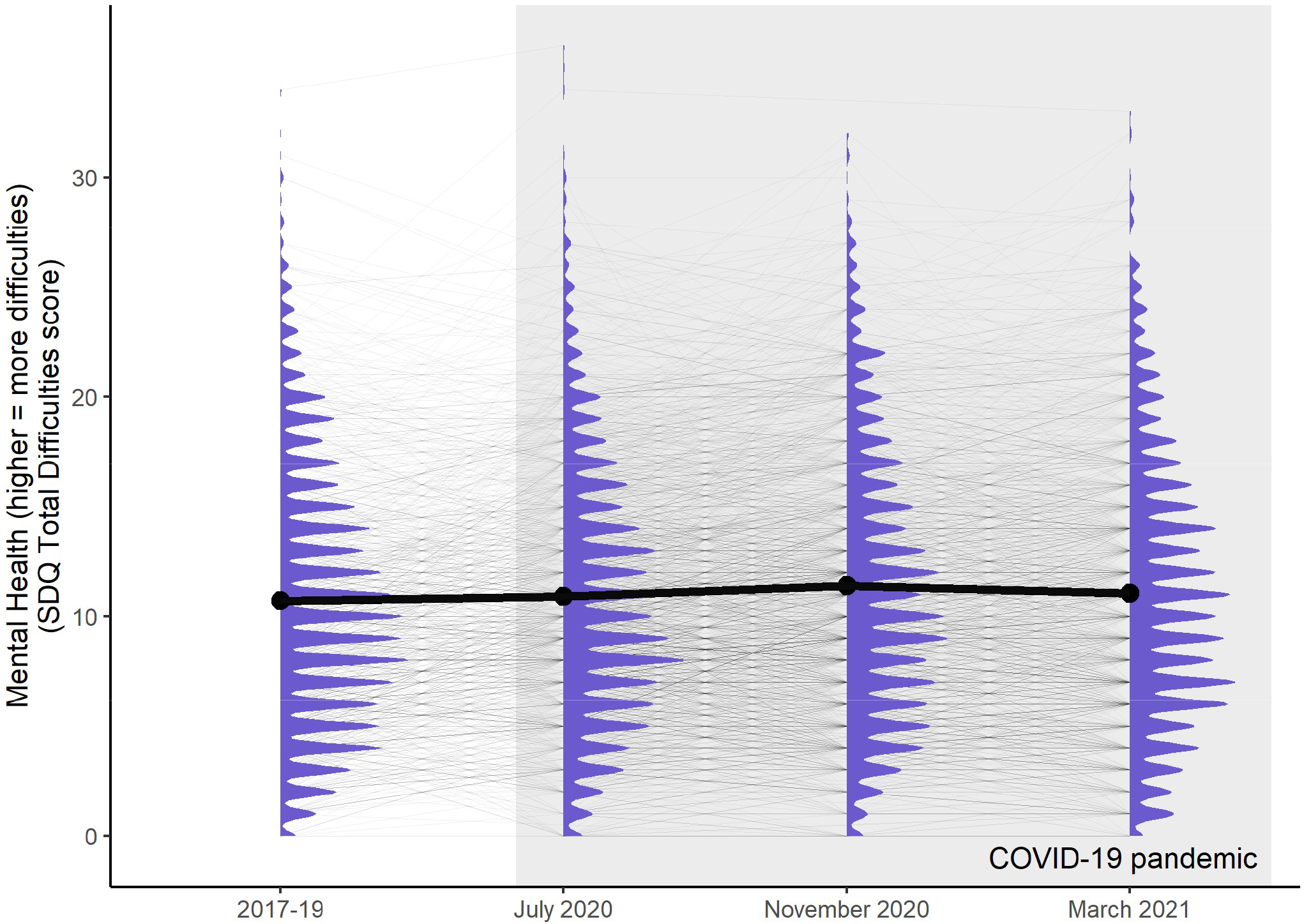
Adolescent SDQ Total Difficulties scores, 2017 to March 2021. For each wave, we plotted the distribution of mental health scores (blue violin plots). The mean of each wave (black) is included to highlight the change in average scores between waves, and individual raw scores are also displayed (grey). The timing of the COVID-19 pandemic is indicated in grey

### Establishing developmental trajectories

To find a best-fit ungrouped LGM for the whole adolescent dataset, we initially fitted an intercept-only model to the whole-cohort Total Difficulties score data (*χ* ^2^ = 182, BIC = 22 133) and compared it to a linear model (*χ* ^2^ = 34·7, BIC = 22 008; LRT *p* < 2 × 10^−16^), which in turn was compared to a quadratic model (*χ* ^2^ = 0·157, BIC = 22 002; LRT *p* = 6 × 10^−7^; for full model fit details see appendix tables A2-3). Based on the significant improvement in model fit, we concluded that the quadratic model (Figure 2) was most appropriate for these data. This supports the previous raw data (Figure 1) showing that mental health followed a quadratic trajectory through the COVID-19 pandemic.

**Figure 2.**
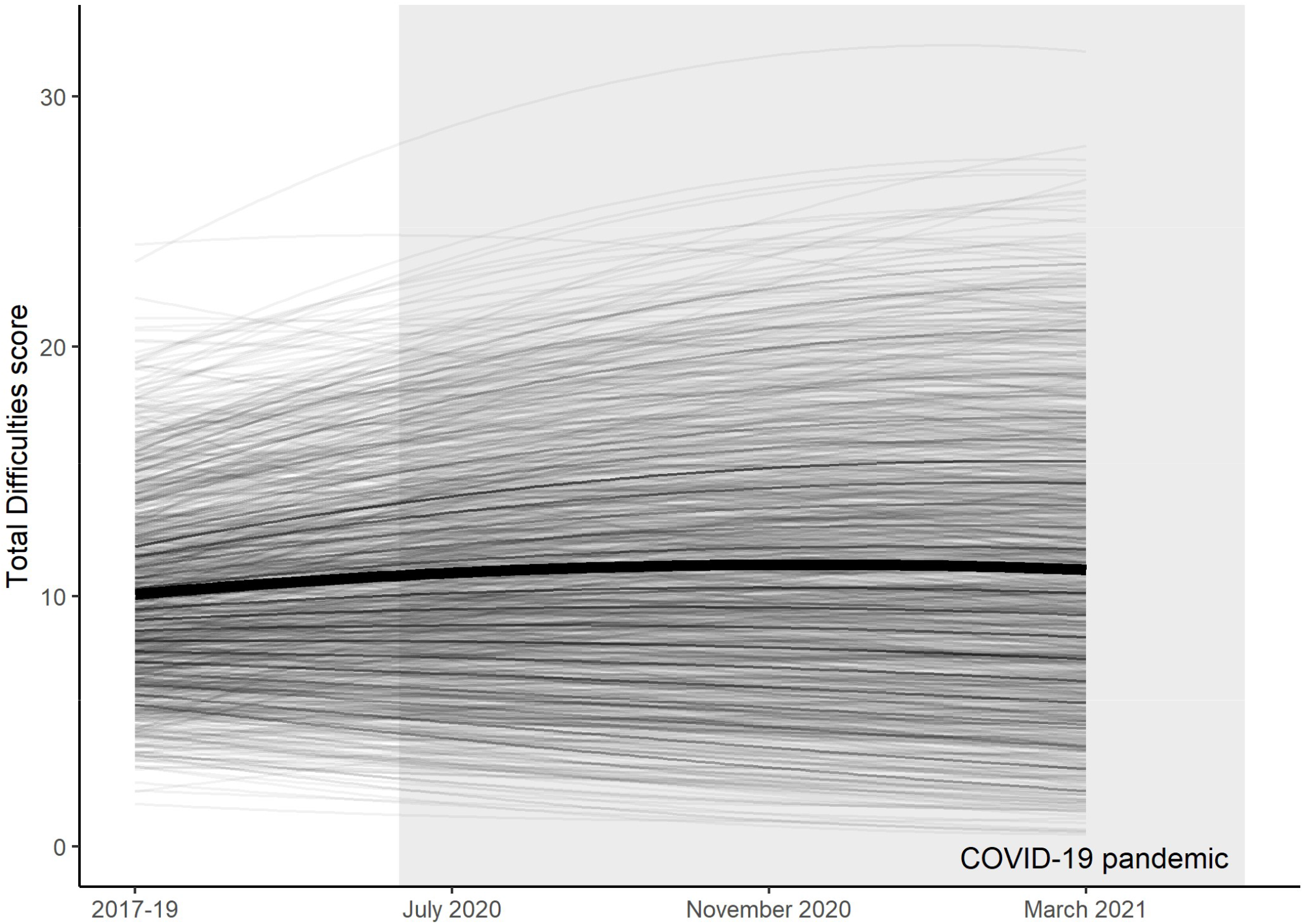
Latent growth curve model (bold) of adolescent SDQ Total Difficulties scores between 2017 and March 2021, based on the Understanding Society dataset. Individual predicted trajectories are also shown, along with the timing of the COVID-19 pandemic in grey.

### Digital inclusion and mental health trajectories

To investigate the impact of computer and internet access on the mental health trajectories plotted in Figure 2, we then fit multi-group LGMs where parameters of interest were selectively constrained to be equal or allowed to vary between digitally excluded and included groups. We fit such multi-group models for both access to a computer and a good internet connection separately.

Access to a computer. For computer access, we found that the modelled linear and quadratic coefficients differed between groups in both models without sociodemographic variables added as control variables (LRT *p* = ·006; p = ·004 respectively; for full details see appendix tables A4-5) and models where control variables were included (LRT *p* = 7 × 10^−4^; *p* = ·004 respectively; for full details see appendix tables A6-7). The group with no computer access has a greatly pronounced increase in mental health symptoms in the early stages of the pandemic, but these returned almost to the level of the group with computer access by COVID-19 wave 8, with (Figure 3, Panel B) or without (Figure 3, Panel A) taking sociodemographic variables into account.

**Figure 3.**
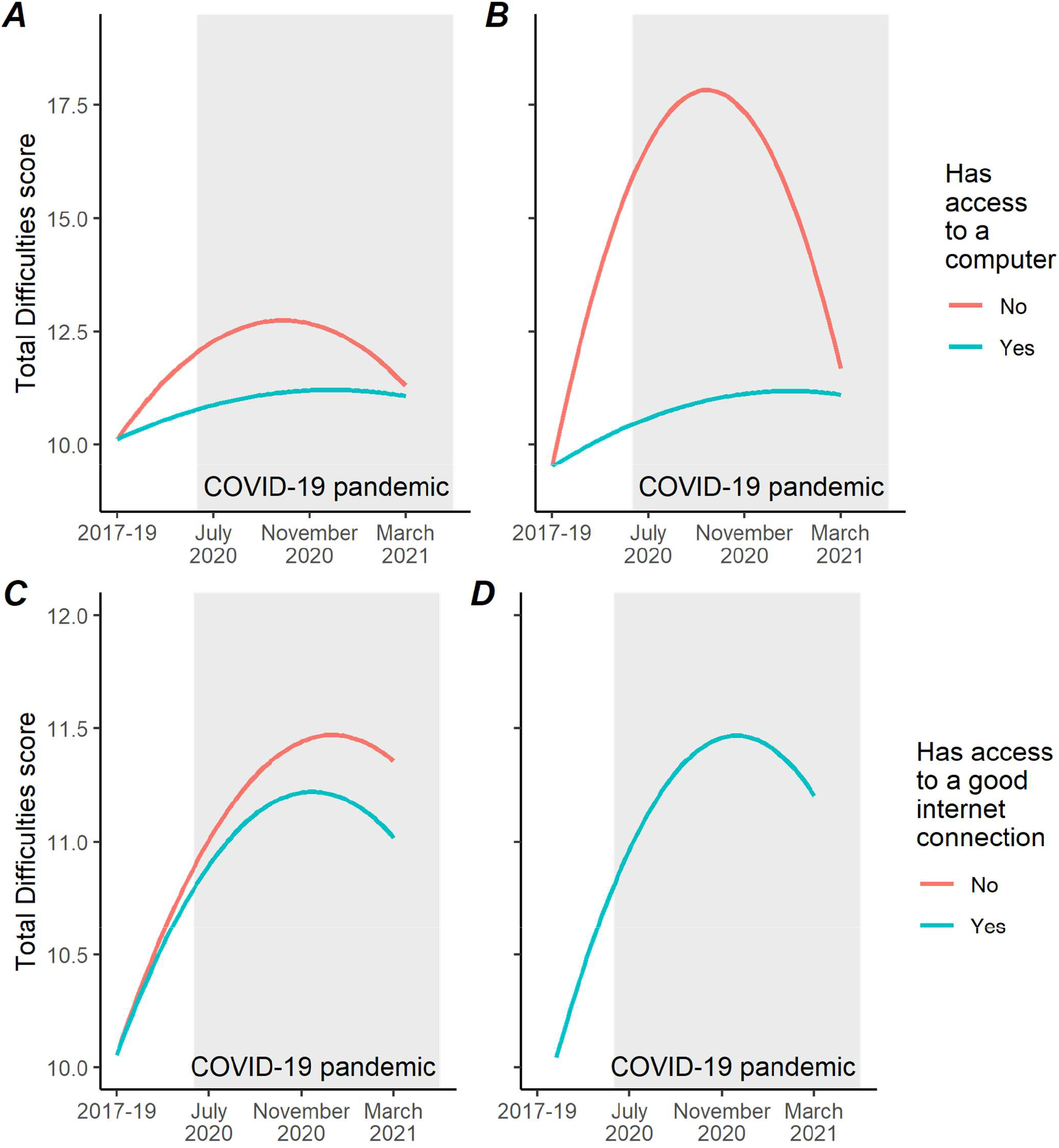
Latent growth curve models of youth SDQ Total Difficulties scores, grouped by each of the two digital inclusion criteria. (A) and (B) show the models grouped by access to a computer - (A) does not include sociodemographic control variables, but does have survey weights applied, while (B) does include the control variables but does not have weights applied (since this process is not robust given the small size of the digitally excluded group). The same apply for (C) and (D), which portray the models grouped by access to a good internet connection – in the latter, the modelled trajectories are identical.

Access to a good internet connection. We applied the same process to test whether access to a good internet connection changed the mental health trajectory across COVID-19. Without accounting for sociodemographic variables (Figure 3, Panel C), the group without good internet access appears to have a slightly more pronounced trajectory, with linear coefficients differing between groups (LRT *p* = ·034; for full details see appendix tables A8-9), however this effect is not significant with control variables accounted for (Figure 3, Panel D; see also appendix tables A10-11), eliminating trajectory differences between groups.

Figure 4 shows the modelled trajectories for the groups without access to a computer and without access to a good internet connection, along with predicted individual trajectories. What is evident in the figure is the small size of the digitally excluded group both for computer access (51 participants), and access to a good internet connection (90 participants).

**Figure 4.**
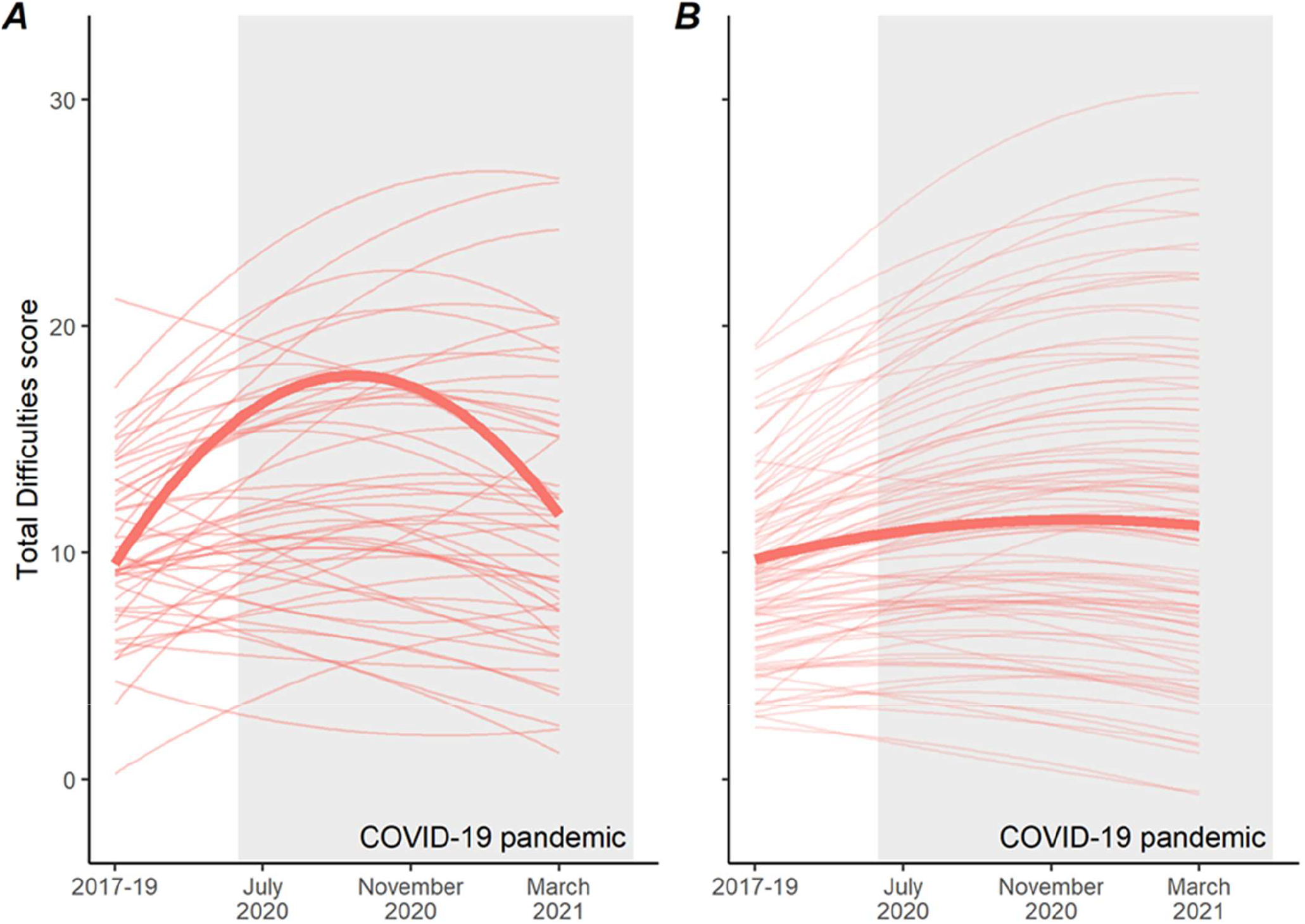
Latent growth curve models of SDQ Total Difficulties scores for those adolescents without access to a computer (A) and without access to a good internet connection (B). Predicted individual trajectories are also shown and demonstrate the size of the group in each case.

### Sensitivity check

To confirm that excluding those participants without assigned longitudinal weights does not substantially affect the results of our analysis, we conducted a sensitivity check including both those with and those without longitudinal weights. These whole-cohort models had only one difference, in that in the “good internet connection” case, the difference in linear coefficients between the groups with and without access became significant (LRT *p*= ·042). Full details of these analyses can be found in the appendix (tables A12-15 and figures A1-2).

## Discussion

In this study we tested whether adolescent longitudinal mental health trajectories during the COVID-19 pandemic were different for adolescents who had experienced digital exclusion during that time to those who had not. When examining all the participating adolescents, we found a small quadratic trend in mental health: symptoms increased from pre-pandemic baselines in 2020 and then decreased in early 2021. The trajectories were more pronounced for those who did not have access to a computer for online schooling, showing a greater increase during 2020 and then a greater decrease in early 2021. In contrast, we found no significant difference between the trajectories for those who had and did not have access to a good internet connection.

To understand these results, it is important to track how the educational and social disruptions experienced by UK adolescents differed across our study waves. In March 2020, UK schools were shut due to the COVID-19 pandemic, except for children of key workers or those who were considered vulnerable to lack of support from school. Furthermore, attendance among these groups was much lower than predicted.^13^ While some schools did reopen before the 2020 summer holidays, attendance was not compulsory and often part-time, especially for older pupils in secondary schools. A full reopening of UK schools only occurred at the beginning of the new school year in September 2020. November 2020 saw a wave of localised restrictions, following which schools were closed again nationally in December 2020 and remained closed until March 2021. Like the Co-Space study, we found mental health symptoms to be worst during times of high COVID-19 restrictions, which would have caused both educational and social disruptions, while mental health recovered to some extent with schools reopening and social restrictions lifting in March 2021.^32^

Our analyses highlighted that those who did not have access to a computer had much worse mental health during times of school closures and social isolation than those who did, which echoes the findings of the English Mental Health of Children and Young People survey follow-ups in 2020 and 2021.^33,34^ Our results were robust even when controlling for sex, age, ethnicity, and household income, which is important as digital exclusion is more likely to co-occur with other adversities. Indeed, both English surveys’ follow-ups demonstrated clustering of various impairments to accessing online schooling in addition to access to a device, including a quiet space to study, support from parents and school, and access to other learning resources.^33,34^ A possible explanation of our results is that digitally excluded adolescents experienced much greater educational and social disruption: a lack of computer access may preclude consistent and active engagement in online schooling and keeping in touch with peers online. These adolescents did not have an effective way to buffer the lack of education or in-person social contact when lockdown measures curtailed their ability to go to school or meet face-to-face, and not having access to such devices can therefore be related to decreased mental health.

There was a clear negative relationship between not having access to a computer for school and mental health, but no evidence for a similar impact of not having access to a “good internet connection”. There are multiple possible explanations for this finding. First, the disruption due to not having a good internet connection (as opposed to having no internet access at all) might not be as severe as not having access to a computer, and the experiences of an adolescent might not have been that different to their peers with a good internet connection. Second, what counts as a “good” internet connection could differ across participants and therefore the measure might have been noisy. As this study highlights the need for further research and policy discussion about digital exclusion considering the COVID-19 pandemic, care needs to be taken to define and discuss digital exclusion and understand its nuances.^35^

Future research should employ a mixed-methods approach to unpick the lockdown experiences of young people and elucidate which aspects of the disruption were most difficult to cope with. It is further possible that the relationship we have identified may extend to other life events beyond the pandemic – therefore, future work may also wish to examine the mental health impacts of digital exclusion beyond the pandemic. It is important to consider these potential effects as digital exclusion frequently co-occurs with other socio-economic disadvantages,^36,37^ with a possibility of associated inequity in mental health risk.

In all, there is a lack of high-quality longitudinal data on digital exclusion and its potential impacts, and further research is necessary. However, our findings combined with those of others^32,34^ suggest that ensuring access to a computer or tablet may be a simple but important intervention should further school closures become necessary.

Our study is limited by the adoption of a narrow operationalisation of digital exclusion, which does not account for the full diversity of affected adolescents. Further, severely digitally excluded adolescents are in the minority in the UK, and therefore, the numbers of adolescent participants in our digitally excluded groups were relatively low. As missing data in the grouping variables could not be imputed, the statistical power of our multigroup LGM analysis was somewhat limited, although our sensitivity checks (see appendix tables A12-15 and figures A1-2) corroborate the study conclusions. In addition, longitudinal sampling weights could not be applied in either case for models incorporating sociodemographic variables, meaning that, for now, those findings cannot be generalised to the whole UK population.

## Research in context

### Evidence before this study

We searched PsycInfo for articles published in English between 1^st^ January 2020 and 1^st^ November 2021 using search terms related to digital access (‘computer’, or ‘laptop’ or ‘digital access’ or digital divide’ or digital inclusion’ or ‘digital exclusion’), mental health (‘psychiatr*’ or ‘mental’ or ‘distress’ or ‘depression’ or ‘anxiety’), COVID (‘covid’ or ‘coronavirus’ or ‘sars-cov-2’), adolescents (‘children’ or ‘adolescents’ or ‘youth’ or ‘child’ or ‘teenager’), and longitudinal analysis (‘trajector*’ or ‘longitudinal’ or ‘latent curve’). While several scholars have theorised the potential negative impact of digital inequalities on psychological well-being, we found no longitudinal studies directly examining whether and how digital exclusion impacted the mental health of adolescents in the context of the pandemic.

### Added value of this study

To our knowledge, this study provides the first longitudinal evidence examining the impact of digital inequalities such as the lack of access to computer for schoolwork on adolescent mental health. As such, our finding that digital exclusion predicts a greater rise in adverse mental health symptoms among UK adolescents during the COVID-19 pandemic is without precedent, and we corroborate hypotheses put forward by scholars as to the negative impact of digital exclusion.

### Implications of all the available evidence

As digitalisation offers crucial opportunities for young people to continue accessing both educational services and social connections, this study indicates that basic levels of digital access can significantly improve mental health outcomes. We suggest that public health officials and policymakers prioritise digital inclusion as part of their pandemic response and recovery plans. Future research is needed to understand and mitigate the adverse mental health impact of digital exclusion, particularly on adolescents from vulnerable and disadvantaged backgrounds.

## Conclusion

We emphasise the urgent need for researchers, public health workers and policy professionals to consider and address digital exclusion as a predictor of adolescent mental health outcomes, especially during the COVID-19 pandemic when much of educational and social life moved onto digital spaces. With digitalisation becoming increasingly widespread in society, ever more services – whether they be educational, social or health-related – are “digital first”, excluding those with little or no access to the devices necessary to engage in these activities.^38^ In the public and scientific conversation that predominantly focuses on the negative impacts of digital technologies on adolescent mental health,^39^ the importance of obtaining basic levels of digital access as a way of supporting adolescent mental health needs to be emphasised more regularly and taken more seriously.

## Supporting information

STROBE checklist for cohort studies

## Data Availability

All data produced are available online at https://osf.io/qhtbj

https://beta.ukdataservice.ac.uk/datacatalogue/doi/?id=6614#14

https://beta.ukdataservice.ac.uk/datacatalogue/doi/?id=8644#10

https://osf.io/qhtbj

## Acknowledgements

TEM was supported by the British Psychological Society Undergraduate Research Assistantship Scheme. SG was supported by the G C Grindley Fund from the School of the Biological Sciences, University of Cambridge. TJF was supported by the NIHR Applied Research Centre; the views expressed are those of the authors, and not necessarily those of the NHS, NIHR or Department of Health and Social Care. AO was supported by the UK Medical Research Council, Economic and Social Research Council, and a College Research Fellowship from Emmanuel College, University of Cambridge.

Understanding Society is an initiative funded by the Economic and Social Research Council and various Government Departments, with scientific leadership by the Institute for Social and Economic Research, University of Essex, and survey delivery by NatCen Social Research and Kantar Public. The research data are distributed by the UK Data Service.

We are grateful for the contributions of Tara Bhagat (University of Cambridge) to synthesising our findings, and of Prof. Rogier Kievit (Donders Institute) to providing assistance with latent growth curve modelling.

This preprint is based on a template created by Brenton M. Wiernik and available at https://osf.io/md9fn/.

## Authors’ contributions

**TEM:** Conceptualisation, methodology, validation, formal analysis, data curation, writing – original draft, writing – review & editing, visualisation, funding acquisition; **SG:** Conceptualisation, methodology, resources, writing – original draft, writing – review & editing, funding acquisition; **EMM:** Methodology, formal analysis, writing – original draft, writing – review & editing; **TJF:** Writing – review & editing; **AO:** Conceptualisation, methodology, validation, resources, writing – original draft, writing – review & editing, supervision, funding acquisition

## Data sharing

All data used in this study are publicly available via the UK Data Service (study numbers 6614 and 8644). R code used for analysis is available at https://osf.io/qhtbj/.

