## Appendix for "Digital exclusion predicts worse mental health among adolescents during COVID-19"

* Author has verified underlying data

**Contents**

Table A1 ... 2
Table A2 ... 2
Table A3 ... 3
Table A4 ... 3
Table A5 ... 4
Table A6 ... 4
Table A7 ... 5
Table A8 ... 6
Table A9 ... 6
Table A10 ... 7
Table A11 ... 8
Table A12 ... 9
Table A13 ... 10
Figure A1 ... 11
Table A14 ... 11
Table A15 ... 12
Figure A2 ... 13

**Table A1**

*Characteristics of participants with, and without, SDQ Total Difficulties scores at each wave (COVID-19 wave 8 and certain other data suppressed to protect participant identities)*

| **Wave** | **All** | **Main study wave 9** | | **COVID-19 wave 4** | | **COVID-19 wave 6** | | **COVID-19 wave 8** | |
|---|---|---|---|---|---|---|---|---|---|
| **SDQ data available** | **All** | **Yes** | **No** | **Yes** | **No** | **Yes** | **No** | **Yes** | **No** |
| **Total** | 1387 | 638 | 749 | 818 | 569 | 836 | 551 | 1386 | 1 |
| **Sex** | | | | | | | | | |
| Male | 656 | 314 | 342 | 372 | 284 | 386 | 270 | ·· | ·· |
| Female | 731 | 324 | 407 | 446 | 285 | 450 | 281 | ·· | ·· |
| Unavailable | 0 | 0 | 0 | 0 | 0 | 0 | 0 | ·· | ·· |
| **Birth year** | | | | | | | | | |
| 2004 | 152 | 129 | 23 | 71 | 81 | 68 | 84 | ·· | ·· |
| 2005 | 209 | 171 | 38 | 149 | 60 | 137 | 72 | ·· | ·· |
| 2006 | 193 | 167 | 26 | 127 | 66 | 124 | 69 | ·· | ·· |
| 2007 | 227 | 130 | 97 | 149 | 78 | 153 | 74 | ·· | ·· |
| 2008 | 213 | 41 | 172 | 138 | 75 | 134 | 79 | ·· | ·· |
| 2009 | 190 | 0 | 190 | 128 | 62 | 129 | 61 | ·· | ·· |
| 2010 | 176 | 0 | 176 | 56 | 120 | 91 | 85 | ·· | ·· |
| 2011 | 27 | 0 | 27 | 0 | 27 | 0 | 27 | ·· | ·· |
| Unavailable | 0 | 0 | 0 | 0 | 0 | 0 | 0 | ·· | ·· |
| **Ethnicity** | | | | | | | | | |
| White British | 973 | 472 | 501 | 590 | 383 | 592 | 381 | ·· | ·· |
| Other White | 32 | 19 | 13 | 16 | 16 | 17 | 15 | ·· | ·· |
| Mixed | 135 | 42 | 93 | 81 | 54 | 84 | 51 | ·· | ·· |
| Asian | 193 | 87 | 106 | 99 | 94 | 109 | 84 | ·· | ·· |
| Black | 34 | 15 | 19 | 20 | 14 | 20 | 14 | ·· | ·· |
| Other | ·· | ·· | ·· | ·· | ·· | ·· | ·· | ·· | ·· |
| Unavailable | ·· | ·· | ·· | ·· | ·· | ·· | ·· | ·· | ·· |
| **Mean household income (x, annual)** | | | | | | | | | |
| x < £20,000 | 90 | 54 | 36 | 65 | 25 | 62 | 28 | ·· | ·· |
| £20,000 ≤ x < £30,000 | 133 | 112 | 21 | 88 | 45 | 93 | 40 | ·· | ·· |
| £30,000 ≤ x < £40,000 | 155 | 134 | 21 | 106 | 49 | 98 | 57 | ·· | ·· |
| £40,000 ≤ x < £50,000 | 149 | 124 | 25 | 102 | 47 | 91 | 58 | ·· | ·· |
| £50,000 ≤ x | 258 | ·· | ·· | 171 | 87 | 167 | 91 | ·· | ·· |
| Unavailable | 602 | ·· | ·· | 286 | 316 | 325 | 277 | ·· | ·· |

**Table A2**

*Model fit statistics for the ungrouped LGM. Decisions on whether to accept or reject each successive model are made using the* p-*value from the relevant pairwise likelihood ratio test (LRT).*

| **Current favoured model** | **Model under test** | **Degrees of freedom (df)** | **Akaike Information Criterion (AIC)** | **Bayesian Information Criterion (BIC)** | $\chi^2$ | **LRT *p*-value** | **Significance (* < 0·05 ** < 0·01 *** < 0·001)** |
|---|---|---|---|---|---|---|---|
| ·· | **No-change** | 8 | 22 101 | 22 133 | 182 | ·· | ·· |
| **No-change** | **Linear** | 5 | 21 961 | 22 008 | 34·7 | $< 2{\cdot}2 \times 10^{-16}$ | *** |
| **Linear** | **Quadratic** | 1 | 21 934 | 22 002 | 0·16 | $5{\cdot}7 \times 10^{-7}$ | *** |

**Table A3**

*Ungrouped model output with survey weights applied*

| | **Estimate(Std.Err.)** | **p** |
|---|---|---|
| **Residual Variances** | | |
| Main study wave 9 | 10·32(1·85) | < 0·001 |
| COVID-19 wave 4 | 6·26(0·63) | < 0·001 |
| COVID-19 wave 6 | 9·03(0·71) | < 0·001 |
| COVID-19 wave 8 | 0·91(1·92) | 0·636 |
| **Latent Intercepts** | | |
| Intercept | 10·10(0·15) | < 0·001 |
| Linear coefficient | 1·12(0·14) | < 0·001 |
| Quadratic coefficient | -0·26(0·04) | < 0·001 |
| **Latent Variances** | | |
| Intercept | 20·72(1·97) | < 0·001 |
| Linear coefficient | 7·32(2·29) | 0·001 |
| Quadratic coefficient | 0·57(0·15) | < 0·001 |
| **Latent Covariances** | | |
| Intercept w/Linear coefficient | -0·10(2·08) | 0·962 |
| Intercept w/Quadratic coefficient | -0·42(0·51) | 0·405 |
| Linear coefficient w/Quadratic coefficient | -1·69(0·53) | 0·001 |
| **Fit Indices** | | |
| $\chi^2$ | 7·88(1) | 0·005 |
| CFI | 1 | |
| RMSEA | 0·07 | |

**Table A4**

*Model fit statistics for model grouped by computer access (no covariates). Model 0 is an unconstrained quadratic model. As before, pairwise LRTs are carried out to determine whether the model with an additional parameter constrained to equality (as demarcated) fits significantly worse than the previous model.*

| **Current favoured model** | **Model under test** | **Additional constraints** | **Degrees of freedom (df)** | **Akaike Information Criterion (AIC)** | **Bayesian Information Criterion (BIC)** | $\chi^2$ | **LRT *p*-value** | **Significance (* < 0·05 ** < 0·01 *** < 0·001)** |
|---|---|---|---|---|---|---|---|---|
| ·· | **0** | ·· | 2 | 15 934 | 16 057 | 5·92 | ·· | ·· |
| **0** | **1** | Residual variances | 6 | 15 929 | 16 033 | 8·88 | 0·54 | ·· |
| **1** | **2** | *y*-intercept | 7 | 15 928 | 16 027 | 9·99 | 0·29 | ·· |
| **2** | ***3*** | *Linear coefficient* | *8* | *15 930* | *16 024* | *14·2* | *0·0064* | ** |
| **2** | ***4*** | *Quadratic coefficient* | *8* | *15 930* | *16 025* | *14·2* | *0·0035* | ** |
| **2** | **5** | *y*-intercept variance | 8 | 15 927 | 16 021 | 11·0 | 0·31 | ·· |
| **5** | **6** | Linear coefficient variance | 9 | 15 925 | 16 015 | 11·1 | 0·89 | ·· |
| **6** | **7** | Quadratic coefficient variance | 10 | 15 924 | 16 009 | 11·8 | 0·40 | ·· |
| **7** | **8** | *y*-intercept/linear coefficient covariance | 11 | 15 922 | 16 002 | 12·1 | 0·57 | ·· |
| **8** | **9** | *y*-intercept/quadratic coefficient covariance | 12 | 15 920 | 15 996 | 12·1 | 0·89 | ·· |
| **9** | ***10*** | *Linear coefficient/quadratic coefficient covariance* | *13* | *15 920* | *15 991* | *14·3* | *0·041* | * |

**Table A5**

*Output for model grouped by computer access (no covariates, survey weights applied)*

| | With access | | Without access | |
|---|---|---|---|---|
| | **Estimate(Std.Err.)** | **p** | **Estimate(Std.Err.)** | **p** |
| **Residual Variances** | | | | |
| Main study wave 9 | 10·46(2·40) | < 0·001 | 10·46(2·40) | < 0·001 |
| COVID-19 wave 4 | 6·27(0·81) | < 0·001 | 6·27(0·81) | < 0·001 |
| COVID-19 wave 6 | 9·06(0·92) | < 0·001 | 9·06(0·92) | < 0·001 |
| COVID-19 wave 8 | 1·00(2·49) | 0·689 | 1·00(2·49) | 0·689 |
| **Latent Intercepts** | | | | |
| Intercept | 10·11(0·19) | < 0·001 | 10·11(0·19) | < 0·001 |
| Linear coefficient | 0·98(0·19) | < 0·001 | 3·05(0·69) | < 0·001 |
| Quadratic coefficient | -0·22(0·05) | < 0·001 | -0·88(0·20) | < 0·001 |
| **Latent Variances** | | | | |
| Intercept | 20·76(2·55) | < 0·001 | 20·76(2·55) | < 0·001 |
| Linear coefficient | 7·07(2·96) | 0·017 | 7·07(2·96) | 0·017 |
| Quadratic coefficient | 0·53(0·19) | 0·005 | 0·53(0·19) | 0·005 |
| **Latent Covariances** | | | | |
| Intercept w/Linear coefficient | -0·23(2·70) | 0·932 | -0·23(2·70) | 0·932 |
| Intercept w/Quadratic coefficient | -0·37(0·66) | 0·57 | -0·37(0·66) | 0·57 |
| Linear coefficient w/Quadratic coefficient | -1·61(0·69) | 0·019 | -1·43(0·70) | 0·041 |
| **Fit Indices** | | | | |
| $\chi^2$ | 39·42(12) | < 0·001 | | |
| CFI | 0·99 | | | |
| RMSEA | 0·07 | | | |

**Table A6**

*Model fit statistics for model grouped by computer access (with covariates). Here the processes as in tables A2 and A4 are combined.*

| **Current favoured model** | **Model under test** | **Additional constraints** | **Degrees of freedom (df)** | **Akaike Information Criterion (AIC)** | **Bayesian Information Criterion (BIC)** | $\chi^2$ | **LRT *p*-value** | **Significance (* < 0·05 ** < 0·01 *** < 0·001)** |
|---|---|---|---|---|---|---|---|---|
| ·· | **No-change** | ·· | 52 | 17 744 | 17 915 | 214 | ·· | ·· |
| **No-change** | **Linear** | ·· | 38 | 17 650 | 17 886 | 91·2 | $< 2{\cdot}2 \times 10^{-16}$ | *** |
| **Linear** | **Quadratic (0)** | ·· | 22 | 17 618 | 17 930 | 27·2 | $1{\cdot}0 \times 10^{-6}$ | *** |
| **0** | **1** | Residual variances | 26 | 17 613 | 17 906 | 30·3 | 0·59 | ·· |
| **1** | **2** | *y*-intercept | 27 | 17 612 | 17 900 | 30·8 | 0·55 | ·· |
| **2** | ***3*** | *Linear coefficient* | *28* | *17 620* | *17 904* | *41·5* | $7{\cdot}2 \times 10^{-4}$ | *** |
| **2** | ***4*** | *Quadratic coefficient* | *28* | *17 621* | *17 905* | *42·7* | *0·0036* | ** |
| **2** | **5** | *y*-intercept variance | 28 | 17 610 | 17 893 | 30·9 | 0·71 | ·· |
| **5** | **6** | Linear coefficient variance | 29 | 17 609 | 17 888 | 32·0 | 0·80 | ·· |
| **6** | **7** | Quadratic coefficient variance | 30 | 17 607 | 17 882 | 32·6 | 0·45 | ·· |
| **7** | **8** | *y*-intercept/linear coefficient covariance | 31 | 17 606 | 17 876 | 33·5 | 0·41 | ·· |
| **8** | **9** | *y*-intercept/quadratic coefficient covariance | 32 | 17 604 | 17 869 | 33·6 | 0·13 | ·· |
| **9** | **10** | Linear coefficient/quadratic coefficient covariance | 33 | 17 603 | 17 864 | 34·7 | 0·29 | ·· |

**Table A7**

*Output for model grouped by computer access (with covariates, no weights applied)*

| | With access | | Without access | |
|---|---|---|---|---|
| | **Estimate(Std.Err.)** | **p** | **Estimate(Std.Err.)** | **p** |
| **Residual Variances** | | | | |
| Sex | 0·25(0·00) | < 0·001 | 0·21(0·03) | < 0·001 |
| Birth year | 0·07(0·00) | < 0·001 | 0·07(0·01) | < 0·001 |
| Dichotomised ethnicity | 0·20(0·01) | < 0·001 | 0·25(0·00) | < 0·001 |
| Mean monthly household income | 0·02(0·00) | < 0·001 | 0·03(0·01) | 0·022 |
| Main study wave 9 | 8·58(4·05) | 0·034 | 8·58(4·05) | 0·034 |
| COVID-19 wave 4 | 10·21(1·35) | < 0·001 | 10·21(1·35) | < 0·001 |
| COVID-19 wave 6 | 8·27(1·35) | < 0·001 | 8·27(1·35) | < 0·001 |
| COVID-19 wave 8 | 4·86(3·79) | 0·2 | 4·86(3·79) | 0·2 |
| **Latent Intercepts** | | | | |
| Intercept | 9·51(0·78) | < 0·001 | 9·51(0·78) | < 0·001 |
| Linear coefficient | 1·35(0·75) | 0·072 | 10·31(3·37) | 0·002 |
| Quadratic coefficient | -0·27(0·20) | 0·182 | -3·20(1·13) | 0·005 |
| **Latent Variances** | | | | |
| Intercept | 21·46(4·39) | < 0·001 | 21·46(4·39) | < 0·001 |
| Linear coefficient | 8·97(5·25) | 0·088 | 8·97(5·25) | 0·088 |
| Quadratic coefficient | 0·38(0·35) | 0·278 | 0·38(0·35) | 0·278 |
| **Latent Covariances** | | | | |
| Intercept w/Linear coefficient | -2·70(4·45) | 0·544 | -2·70(4·45) | 0·544 |
| Intercept w/Quadratic coefficient | 0·27(1·04) | 0·796 | 0·27(1·04) | 0·796 |
| Linear coefficient w/Quadratic coefficient | -1·67(1·26) | 0·185 | -1·67(1·26) | 0·185 |
| **Fit Indices** | | | | |
| $\chi^2$ | 34·72 | | | |
| CFI | 1 | | | |
| RMSEA | 0·01 | | | |
| Scaled $\chi^2$ | 36·05(33) | 0·328 | | |

**Table A8**

*Model fit statistics for model grouped by good internet connection access (no covariates). Processes identical to those in table A6.*

| Current favoured model | Model under test | Additional constraints | Degrees of freedom (df) | Akaike Information Criterion (AIC) | Bayesian Information Criterion (BIC) | $\chi^2$ | LRT *p*-value | Significance (* < 0·05 ** < 0·01 *** < 0·001) |
|---|---|---|---|---|---|---|---|---|
| ·· | **0** | ·· | 2 | 15 926 | 16 048 | 1·78 | ·· | ·· |
| **0** | **1** | Residual variances | 6 | 15 921 | 16 025 | 5·51 | 0·56 | ·· |
| **1** | **2** | *y*-intercept | 7 | 15 920 | 16 019 | 5·76 | 0·61 | ·· |
| **2** | ***3*** | *Linear coefficient* | *8* | *15 921* | *16 016* | *9·36* | *0·034* | * |
| **2** | **4** | Quadratic coefficient | 8 | 15 920 | 16 015 | 8·39 | 0·080 | ·· |
| **4** | **5** | *y*-intercept variance | 9 | 15 919 | 16 009 | 8·97 | 0·44 | ·· |
| **5** | **6** | Linear coefficient variance | 10 | 15 917 | 16 002 | 8·97 | 0·99 | ·· |
| **6** | ***7*** | *Quadratic coefficient variance* | *11* | *15 918* | *15 998* | *11·8* | *0·044* | * |
| **6** | **8** | *y*-intercept/linear coefficient covariance | 11 | 15 915 | 15 995 | 9·18 | 0·62 | ·· |
| **8** | **9** | *y*-intercept/quadratic coefficient covariance | 12 | 15 915 | 15 991 | 11·5 | 0·12 | ·· |
| **9** | ***10*** | *Linear coefficient/quadratic coefficient covariance* | *13* | *15 922* | *15 993* | *19·9* | *$5{\cdot}9 \times 10^{-4}$* | *** |

**Table A9**

*Output for model grouped by good internet connection access (no covariates, survey weights applied)*

| | **With access** | | **Without access** | |
|---|---|---|---|---|
| | **Estimate(Std.Err.)** | **p** | **Estimate(Std.Err.)** | **p** |
| **Residual Variances** | | | | |
| Main study wave 9 | 10·74(2·40) | < 0·001 | 10·74(2·40) | < 0·001 |
| COVID-19 wave 4 | 6·32(0·81) | < 0·001 | 6·32(0·81) | < 0·001 |
| COVID-19 wave 6 | 8·99(0·92) | < 0·001 | 8·99(0·92) | < 0·001 |
| COVID-19 wave 8 | 0·78(2·49) | 0·754 | 0·78(2·49) | 0·754 |
| **Latent Intercepts** | | | | |
| Intercept | 10·05(0·19) | < 0·001 | 10·05(0·19) | < 0·001 |
| Quadratic coefficient | -0·26(0·05) | < 0·001 | -0·26(0·05) | < 0·001 |
| Linear coefficient | 1·10(0·18) | < 0·001 | 1·22(0·28) | < 0·001 |
| **Latent Variances** | | | | |
| Intercept | 20·62(2·55) | < 0·001 | 20·62(2·55) | < 0·001 |
| Linear coefficient | 6·82(2·96) | 0·021 | 6·82(2·96) | 0·021 |
| Quadratic coefficient | 0·56(0·19) | 0·003 | 0·15(0·23) | 0·508 |
| **Latent Covariances** | | | | |
| Intercept w/Linear coefficient | -0·01(2·69) | 0·998 | -0·01(2·69) | 0·998 |
| Intercept w/Quadratic coefficient | -0·44(0·65) | 0·5 | -0·44(0·65) | 0·5 |
| Linear coefficient w/Quadratic coefficient | -1·62(0·69) | 0·019 | -0·75(0·73) | 0·302 |
| **Fit Indices** | | | | |
| $\chi^2$ | 49·42(12) | < 0·001 | | |
| CFI | 0·98 | | | |
| RMSEA | 0·09 | | | |

**Table A10**

*Model fit statistics for model grouped by good internet connection access (with covariates). Processes identical to those in tables A6 and A8.*

| **Current favoured model** | **Model under test** | **Additional constraints** | **Degrees of freedom (df)** | **Akaike Information Criterion (AIC)** | **Bayesian Information Criterion (BIC)** | $\chi^2$ | **LRT *p*-value** | **Significance (* < 0·05 ** < 0·01 *** < 0·001)** |
|---|---|---|---|---|---|---|---|---|
| ·· | **No-change** | ·· | 52 | 17 771 | 17 941 | 206 | ·· | ·· |
| **No-change** | **Linear** | ·· | 38 | 17 672 | 17 908 | 78·6 | $< 2·2 \times 10^{-16}$ | *** |
| **Linear** | **Quadratic (0)** | ·· | 22 | 17 652 | 17 964 | 26·7 | $6·1 \times 10^{-5}$ | *** |
| **0** | **1** | Residual variances | 26 | 17 647 | 17 940 | 29·8 | 0·65 | ·· |
| **1** | **2** | *y*-intercept | 27 | 17 646 | 17 934 | 30·8 | 0·26 | ·· |
| **2** | **3** | Linear coefficient | 28 | 17 645 | 17 929 | 31·7 | 0·33 | ·· |
| **3** | **4** | Quadratic coefficient | 29 | 17 644 | 17 923 | 32·6 | 0·47 | ·· |
| **4** | **5** | *y*-intercept variance | 30 | 17 642 | 17 917 | 33·2 | 0·55 | ·· |
| **5** | **6** | Linear coefficient variance | 31 | 17 640 | 17 910 | 33·3 | 0·78 | ·· |
| **6** | **7** | Quadratic coefficient variance | 32 | 17 642 | 17 907 | 36·7 | 0·062 | ·· |
| **7** | **8** | *y*-intercept/linear coefficient covariance | 33 | 17 640 | 17 900 | 36·7 | 0·99 | ·· |
| **8** | ***9*** | *y-intercept/quadratic coefficient covariance* | *34* | *17 646* | *17 902* | *45·3* | *0·015* | * |
| **8** | **10** | Linear coefficient/quadratic coefficient covariance | 34 | 17 641 | 17 897 | 40·3 | 0·22 | ·· |

**Table A11**

*Output for model grouped by good internet connection access (with covariates, no weights applied)*

| | **With access** | | **Without access** | |
|---|---|---|---|---|
| | **Estimate(Std.Err.)** | **p** | **Estimate(Std.Err.)** | **p** |
| **Residual Variances** | | | | |
| Sex | 0·25(0·00) | < 0·001 | 0·23(0·01) | < 0·001 |
| Birth year | 0·07(0·00) | < 0·001 | 0·06(0·01) | < 0·001 |
| Dichotomised ethnicity | 0·20(0·01) | < 0·001 | 0·22(0·02) | < 0·001 |
| Mean monthly household income | 0·02(0·00) | < 0·001 | 0·02(0·01) | 0·028 |
| Main study wave 9 | 10·08(4·12) | 0·014 | 10·08(4·12) | 0·014 |
| COVID-19 wave 4 | 10·12(1·34) | < 0·001 | 10·12(1·34) | < 0·001 |
| COVID-19 wave 6 | 8·47(1·34) | < 0·001 | 8·47(1·34) | < 0·001 |
| COVID-19 wave 8 | 5·20(3·80) | 0·171 | 5·20(3·80) | 0·171 |
| **Latent Intercepts** | | | | |
| Intercept | 9·72(0·78) | < 0·001 | 9·72(0·78) | < 0·001 |
| Linear coefficient | 1·62(0·74) | 0·029 | 1·62(0·74) | 0·029 |
| Quadratic coefficient | -0·38(0·20) | 0·063 | -0·38(0·20) | 0·063 |
| **Latent Variances** | | | | |
| Intercept | 20·51(4·48) | < 0·001 | 20·51(4·48) | < 0·001 |
| Linear coefficient | 7·47(5·45) | 0·171 | 7·47(5·45) | 0·171 |
| Quadratic coefficient | 0·28(0·37) | 0·443 | 0·28(0·37) | 0·443 |
| **Latent Covariances** | | | | |
| Intercept w/Linear coefficient | -1·56(4·58) | 0·733 | -1·56(4·58) | 0·733 |
| Linear coefficient w/Quadratic coefficient | -1·31(1·32) | 0·323 | -1·31(1·32) | 0·323 |
| Intercept w/Quadratic coefficient | -0·05(1·08) | 0·962 | 0·75(1·15) | 0·511 |
| **Fit Indices** | | | | |
| $\chi^2$ | 40·29 | | | |
| CFI | 1 | | | |
| RMSEA | 0·02 | | | |
| Scaled $\chi^2$ | 37·28(34) | 0·321 | | |

**Table A12**

*Sensitivity check: model fit statistics for model grouped by computer access (no covariates, participants without CV8 longitudinal weight NOT excluded). Processes identical to those in tables A6, A8 and A10.*

| **Current favoured model** | **Model under test** | **Additional constraints** | **Degrees of freedom (df)** | **Akaike Information Criterion (AIC)** | **Bayesian Information Criterion (BIC)** | $\chi^2$ | **LRT *p*-value** | **Significance (* < 0·05 ** < 0·01 *** < 0·001)** |
|---|---|---|---|---|---|---|---|---|
| ·· | **No-change** | ·· | 52 | 26 311 | 26 500 | 299 | ·· | ·· |
| **No-change** | **Linear** | ·· | 38 | 26 177 | 26 440 | 137 | < 2·2 × $10^{-16}$ | *** |
| **Linear** | **Quadratic (0)** | ·· | 22 | 26 114 | 26 461 | 41·9 | 2·5 × $10^{-12}$ | *** |
| **0** | **1** | Residual variances | 26 | 26 110 | 26 437 | 46·6 | 0·32 | ·· |
| **1** | **2** | *y*-intercept | 27 | 26 110 | 26 431 | 48·4 | 0·31 | ·· |
| **2** | ***3*** | *Linear coefficient* | *28* | *26 115* | *26 431* | *55·5* | *0·0032* | ** |
| **2** | ***4*** | *Quadratic coefficient* | *28* | *26 115* | *26 431* | *55·7* | *0·0096* | ** |
| **2** | **5** | *y*-intercept variance | 28 | 26 109 | 26 424 | 48·7 | 0·64 | ·· |
| **5** | **6** | Linear coefficient variance | 29 | 26 109 | 26 419 | 50·8 | 0·28 | ·· |
| **6** | **7** | Quadratic coefficient variance | 30 | 26 108 | 26 413 | 52·5 | 0·063 | ·· |
| **7** | **8** | *y*-intercept/linear coefficient covariance | 31 | 26 107 | 26 407 | 53·2 | 0·45 | ·· |
| **8** | **9** | *y*-intercept/quadratic coefficient covariance | 32 | 26 105 | 26 400 | 53·3 | 0·75 | ·· |
| **9** | **10** | Linear coefficient/quadratic coefficient covariance | 33 | 26 104 | 26 393 | 53·9 | 0·12 | ·· |

**Table A13**

*Output for sensitivity check computer model with survey weights applied*

| | With access | | Without access | |
|---|---|---|---|---|
| | **Estimate(Std.Err.)** | **p** | **Estimate(Std.Err.)** | **p** |
| **Residual Variances** | | | | |
| Sex | 0·25(0·00) | < 0·001 | 0·25(0·00) | < 0·001 |
| Birth year | 0·09(0·00) | < 0·001 | 0·10(0·01) | < 0·001 |
| Dichotomised ethnicity | 0·22(0·00) | < 0·001 | 0·25(0·00) | < 0·001 |
| Mean monthly household income | 0·02(0·00) | < 0·001 | 0·02(0·01) | 0·015 |
| Main study wave 9 | 11·05(3·62) | 0·002 | 11·05(3·62) | 0·002 |
| COVID-19 wave 4 | 9·93(1·08) | < 0·001 | 9·93(1·08) | < 0·001 |
| COVID-19 wave 6 | 8·28(1·10) | < 0·001 | 8·28(1·10) | < 0·001 |
| COVID-19 wave 8 | 4·71(3·44) | 0·171 | 4·71(3·44) | 0·171 |
| **Latent Intercepts** | | | | |
| Intercept | 10·20(0·64) | < 0·001 | 10·20(0·64) | < 0·001 |
| Linear coefficient | 1·20(0·64) | 0·061 | 6·33(2·26) | 0·005 |
| Quadratic coefficient | -0·22(0·19) | 0·225 | -2·13(0·86) | 0·013 |
| **Latent Variances** | | | | |
| Intercept | 21·87(3·73) | < 0·001 | 21·87(3·73) | < 0·001 |
| Linear coefficient | 7·61(4·63) | 0·1 | 7·61(4·63) | 0·1 |
| Quadratic coefficient | 0·33(0·33) | 0·329 | 0·33(0·33) | 0·329 |
| **Latent Covariances** | | | | |
| Intercept w/Linear coefficient | -1·39(3·84) | 0·718 | -1·39(3·84) | 0·718 |
| Intercept w/Quadratic coefficient | -0·05(0·93) | 0·955 | -0·05(0·93) | 0·955 |
| Linear coefficient w/Quadratic coefficient | -1·39(1·15) | 0·226 | -1·39(1·15) | 0·226 |
| **Fit Indices** | | | | |
| $\chi^2$ | 53.9 | | | |
| CFI | 0.99 | | | |
| RMSEA | 0.03 | | | |
| Scaled $\chi^2$ | 53.96(33) | 0.012 | | |

**Figure A1**

*Sensitivity check: latent growth curve model of youth SDQ Total Difficulties scores, grouped by computer access, including sociodemographic control variables and without excluding participants with no longitudinal weight for COVID-19 wave 8.*

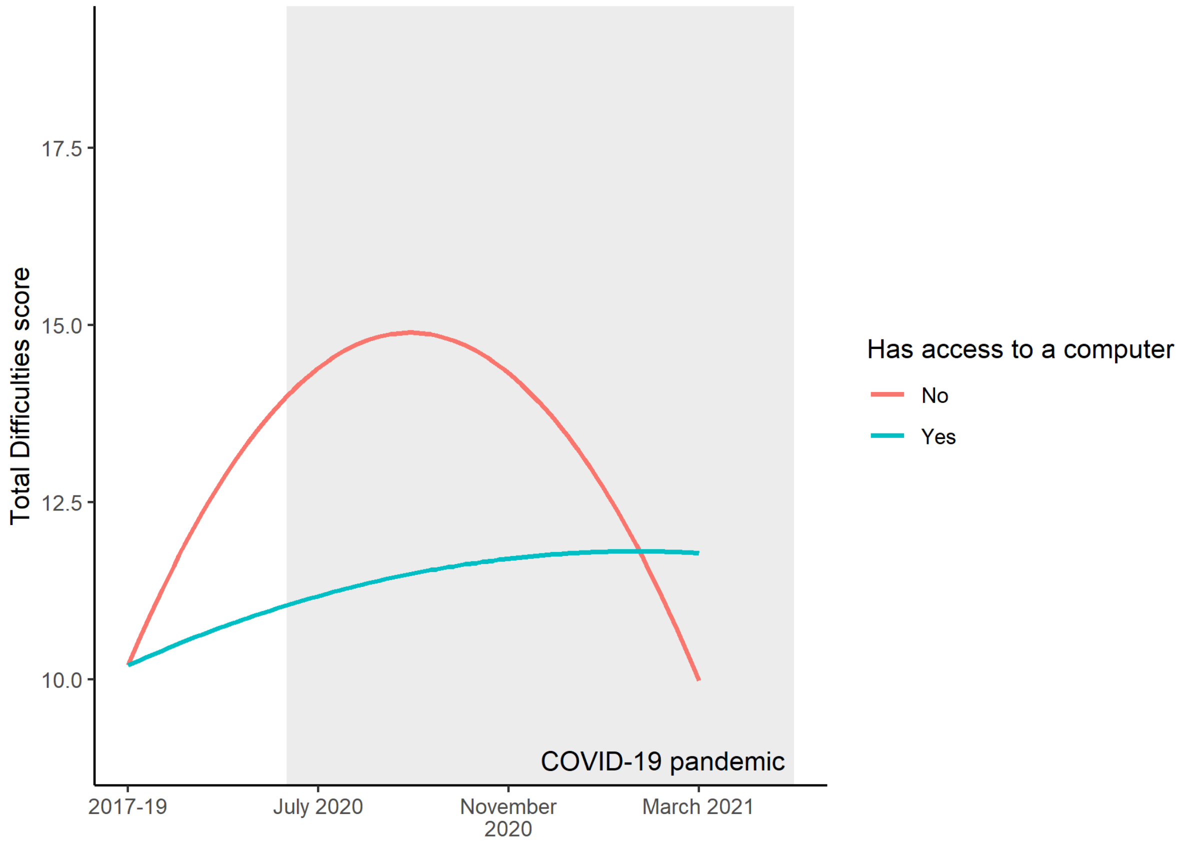


**Table A14**

*Sensitivity check: model fit statistics for model grouped by access to a good internet connection (no covariates, participants without CV8 longitudinal weight NOT excluded). Processes identical to those in tables A6, A8, A10 and A12.*

| **Current favoured model** | **Model under test** | **Additional constraints** | **Degrees of freedom (df)** | **Akaike Information Criterion (AIC)** | **Bayesian Information Criterion (BIC)** | $\chi^2$ | **LRT *p*-value** | **Significance (* < 0·05 ** < 0·01 *** < 0·001)** |
|---|---|---|---|---|---|---|---|---|
| ·· | **No-change** | ·· | 52 | 26 324 | 26 514 | 289 | ·· | ·· |
| **No-change** | **Linear** | ·· | 38 | 26 191 | 26 454 | 127 | $< 2{\cdot}2 \times 10^{-16}$ | *** |
| **Linear** | **Quadratic (0)** | ·· | 22 | 26 133 | 26 480 | 37·2 | $4{\cdot}3 \times 10^{-11}$ | *** |
| **0** | **1** | Residual variances | 26 | 26 128 | 26 455 | 41·2 | 0·53 | ·· |
| **1** | **2** | *y*-intercept | 27 | 26 126 | 26 447 | 41·2 | 0·96 | ·· |
| **2** | ***3*** | *Linear coefficient* | *28* | *26 129* | *26 445* | *45·8* | *0·042* | * |
| **2** | **4** | Quadratic coefficient | 28 | 26 129 | 26 445 | 45·7 | 0·089 | ·· |
| **4** | **5** | *y*-intercept variance | 29 | 26 127 | 26 437 | 45·8 | 0·74 | ·· |
| **5** | **6** | Linear coefficient variance | 30 | 26 125 | 26 430 | 45·8 | 0·89 | ·· |
| **6** | **7** | Quadratic coefficient variance | 31 | 26 127 | 26 426 | 49·2 | 0·067 | ·· |
| **7** | **8** | *y*-intercept/linear coefficient covariance | 32 | 26 125 | 26 419 | 49·6 | 0·55 | ·· |
| **8** | ***9*** | *y-intercept/quadratic coefficient covariance* | *33* | *26 130* | *26 419* | *56·5* | *0·039* | * |
| **8** | **10** | Linear coefficient/quadratic coefficient covariance | 33 | 26 126 | 26 415 | 52·4 | 0·28 | ·· |

**Table A15**

*Output for sensitivity check internet model with survey weights applied*

| | With access | | Without access | |
|---|---|---|---|---|
| | **Estimate(Std.Err.)** | **p** | **Estimate(Std.Err.)** | **p** |
| **Residual Variances** | | | | |
| Sex | 0·25(0·00) | < 0·001 | 0·25(0·00) | < 0·001 |
| Birth year | 0·09(0·00) | < 0·001 | 0·08(0·01) | < 0·001 |
| Dichotomised ethnicity | 0·22(0·00) | < 0·001 | 0·23(0·01) | < 0·001 |
| Mean monthly household income | 0·02(0·00) | < 0·001 | 0·02(0·01) | 0·003 |
| Main study wave 9 | 11·69(3·65) | 0·001 | 11·69(3·65) | 0·001 |
| COVID-19 wave 4 | 9·94(1·07) | < 0·001 | 9·94(1·07) | < 0·001 |
| COVID-19 wave 6 | 8·30(1·09) | < 0·001 | 8·30(1·09) | < 0·001 |
| COVID-19 wave 8 | 5·42(3·47) | 0·119 | 5·42(3·47) | 0·119 |
| **Latent Intercepts** | | | | |
| Intercept | 10·32(0·64) | < 0·001 | 10·32(0·64) | < 0·001 |
| Quadratic coefficient | -0·33(0·18) | 0·072 | -0·33(0·18) | 0·072 |
| Linear coefficient | 1·44(0·63) | 0·022 | 1·96(0·85) | 0·021 |
| **Latent Variances** | | | | |
| Intercept | 21·40(3·76) | < 0·001 | 21·40(3·76) | < 0·001 |
| Linear coefficient | 7·07(4·72) | 0·135 | 7·07(4·72) | 0·135 |
| Quadratic coefficient | 0·26(0·35) | 0·451 | 0·26(0·35) | 0·451 |
| **Latent Covariances** | | | | |
| Intercept w/Linear coefficient | -0·94(3·90) | 0·809 | -0·94(3·90) | 0·809 |
| Linear coefficient w/Quadratic coefficient | -1·22(1·18) | 0·302 | -1·22(1·18) | 0·302 |
| Intercept w/Quadratic coefficient | -0·22(0·95) | 0·818 | 0·45(1·02) | 0·656 |
| **Fit Indices** | | | | |
| $\chi^2$ | 52·44 | | | |
| CFI | 0·99 | | | |
| RMSEA | 0·03 | | | |
| Scaled $\chi^2$ | 48·77(33) | 0·038 | | |

**Figure A2**

*Sensitivity check: latent growth curve model of youth SDQ Total Difficulties scores, grouped by access to a good internet connection, including sociodemographic control variables and without excluding participants with no longitudinal weight for COVID-19 wave 8.*

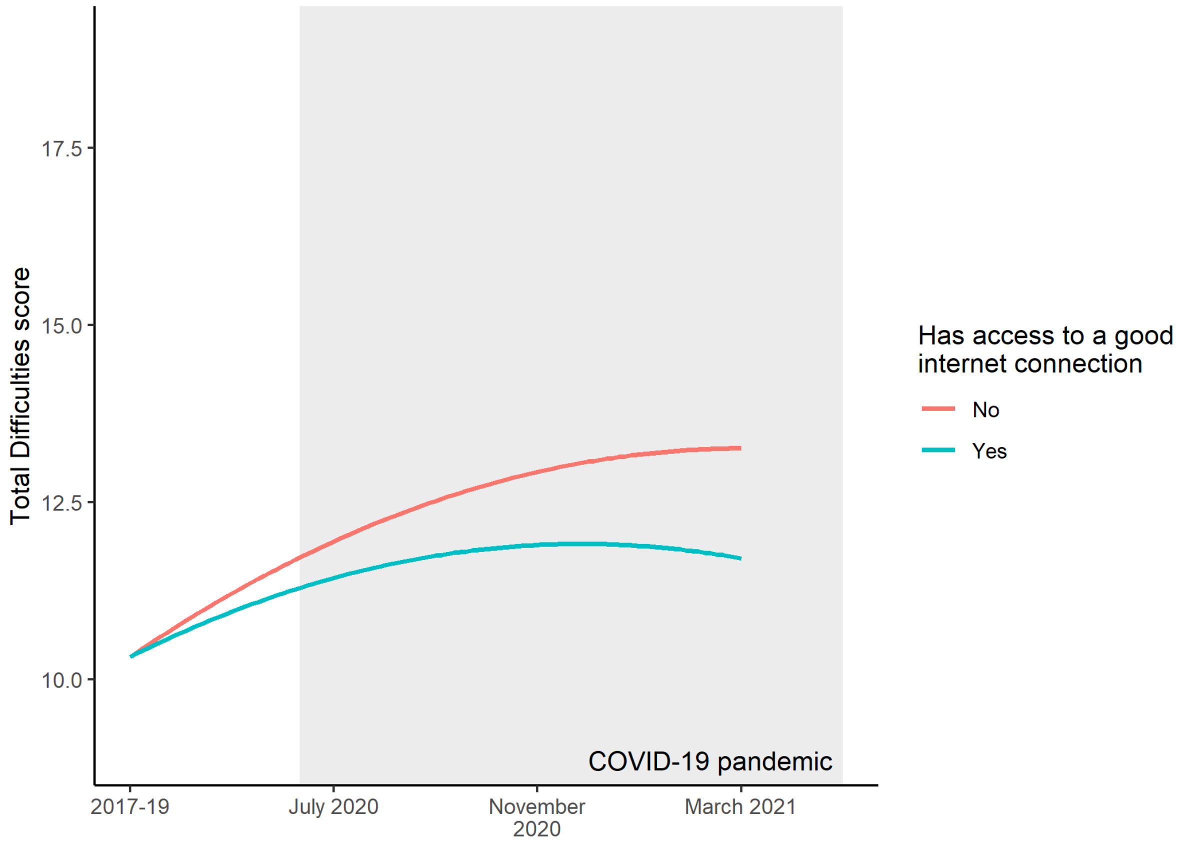
